# COMET-LF: A Compartmental Model of Dynamics of Infection, Disease, and Elimination Strategies for Lymphatic Filariasis

**DOI:** 10.1101/2024.09.27.24314480

**Authors:** Indrajit Ghosh, Suchita Nath-Sain, Shoummo Sen Gupta, Chhavi Pant Joshi, Tanu Jain, Swaminathan Subramanian, Souvik Banerjee, Mithun Kumar Mitra

**Affiliations:** National Disease Modelling Consortium, Indian Institute of Technology Bombay, Mumbai, Maharashtra, India; Koita Centre for Digital Health, Indian Institute of Technology Bombay, Mumbai, Maharashtra, India; National Center for Vector Borne Diseases Control, New Delhi, India; Consultant, National Disease Modelling Consortium, Formerly at ICMR-Vector Control Research Centre, Puducherry, India; Department of Economics, Indian Institute of Technology Bombay, Mumbai, Maharashtra, India; Department of Physics, Indian Institute of Technology Bombay, Mumbai, Maharashtra, India

## Abstract

Lymphatic filariasis (LF) is a mosquito-borne neglected tropical disease (NTD) caused by filarial worms. India accounted for 55% of the global population at risk of LF in 2021. The World Health Organization (WHO) has targeted LF elimination by 2030; however, India aims to achieve LF elimination prior to the global WHO NTD target. Mathematical models are useful tools to evaluate and guide elimination strategies. We propose a new compartmental model—COmpartmental Modelling of Elimination strategies and Transmission of Lymphatic Filariasis (COMET-LF)—to assess the impact of mass drug administration (MDA) on LF elimination. Our model incorporates drug efficacy data from a clinical trial and generates estimates of disease (lymphoedema and hydrocele) prevalence. The model is calibrated to publicly available microfilaria (Mf) and disease prevalence data (2008-2013) from Bihar, India. Predictions of the number of MDA rounds needed for achieving the elimination threshold were generated for various endemic scenarios. The projected estimates were compared with established micro- (LYMFASIM) and macro- (EPIFIL) simulation models for LF transmission. Disease burden estimates and the impact of MDA on disease burden were generated using COMET-LF for different endemic scenarios. Our simulations suggest that the disease burden reduces over much longer timescales - 20 years for a reduction of 8%-11.5% following 5 rounds of MDA. We extended COMET-LF to a meta-population model to investigate the role of migration among neighbouring regions on elimination and resurgence probabilities. We found that high Mf prevalence in the spatial neighbourhood can increase the number of required MDA rounds for elimination up to 3 additional rounds for the two-drug regimen. Furthermore, we assess the impact of migration on the resurgence probability in a non-endemic region which is spatially adjacent to a high-Mf prevalence region and show that there is a significant risk of resurgence if Mf prevalence exceeds 5%. Our model can be easily tailored to specific blocks and districts to guide programmatic intervention for disease management and LF elimination.

**Author summary:** Lymphatic filariasis (LF) commonly occurs in tropical regions and is transmitted to humans by mosquitoes infected with larvae of parasitic roundworms. Some patients develop external symptoms including swollen limbs/male genitals that develop from damage to lymph nodes. Others do not develop external symptoms but may transmit the disease to non-infected humans through mosquito bites. LF causes physical disability, disfigurement and mental suffering. India has more than half of the global population at risk of developing LF. Currently, medications that kill the parasites are given yearly to the population at risk. A better understanding of the disease transmission and control measures is important to meet the 2030 elimination target set by the World Health Organization. We developed a new mathematical model (COMET-LF) that takes into account India-specific disease information for more accurate predictions. To validate our model, we compared the predictions with those from established models. COMET-LF can predict the number of years the drug has to be administered to stop LF transmission and the effect of drugs on disease prevalence. COMET-LF also shows that infected patients migrating from neighboring regions can increase transmission to regions where LF is under control. Notably, our model can help policy makers plan targeted control measures for specific regions.

## 1 Introduction

Lymphatic filariasis (LF) is a debilitating neglected tropical disease that affected over 50 million people worldwide in 2018 [1]. In 2000, the World Health Organization (WHO) launched the Global Programme to Eliminate Lymphatic Filariasis (GPELF). Since then, the programme has made significant progress in achieving elimination in 19 of the 72 countries endemic for LF [2]. In 2020, WHO set a goal to eliminate LF by 2030 [3]. India, accounting 55% of the global population at risk of LF in 2021 [4], has committed to eliminating LF before the global target. A total of 6,19,426 lymphoedema and 1,26,906 hydrocele cases were reported in 2023 with the highest burden from 8 states including Bihar (lymphoedema, 25%; hydrocele, 18%), Uttar Pradesh (15%; 19%), and Jharkhand (9%; 33%) [5]. Furthermore, the programme success varied among different endemic regions in India. Therefore, a region-specific understanding of the disease is needed for geographically tailored strategies to achieve the elimination target.

LF is a mosquito-borne disease caused by the filarial worms *Wuchereria bancrofti, Brugia malayi*, and *Brugia timori* [6]. Of these, *W. bancrofti* causes approximately 99.4% of LF cases in India [7]. The filarial worm has a complex life cycle involving mosquitoes and humans (Fig. 1). *Culex quinquefasciatus* is the major vector in India [8]. When a female mosquito feeds on an infected person, it ingests microfilariae (Mf), which are immature larvae that circulate in the bloodstream. The Mf develop into L3 larvae in ≈ 10-12 days inside the mosquito. During a subsequent blood meal the infected mosquito transmits the L3 larvae, which mature into adult worms in the human lymphatic vessels. Subsequently, the adult worms produce Mf [9]. LF infection causes asymptomatic, acute, and chronic conditions [10]. Although most of the LF infections are asymptomatic, these individuals can transmit Mf to mosquitoes and, therefore, contribute to disease transmission. Chronic conditions of LF infection include lymphoedema (swelling of lymph nodes), elephantiasis (an advanced form of lymphoedema causing swelling of the limbs), and hydrocele (swelling of scrotum). Such conditions often lead to social stigma, worse mental health, and poor labour market opportunities and outcomes [3, 11].

**Fig 1.**
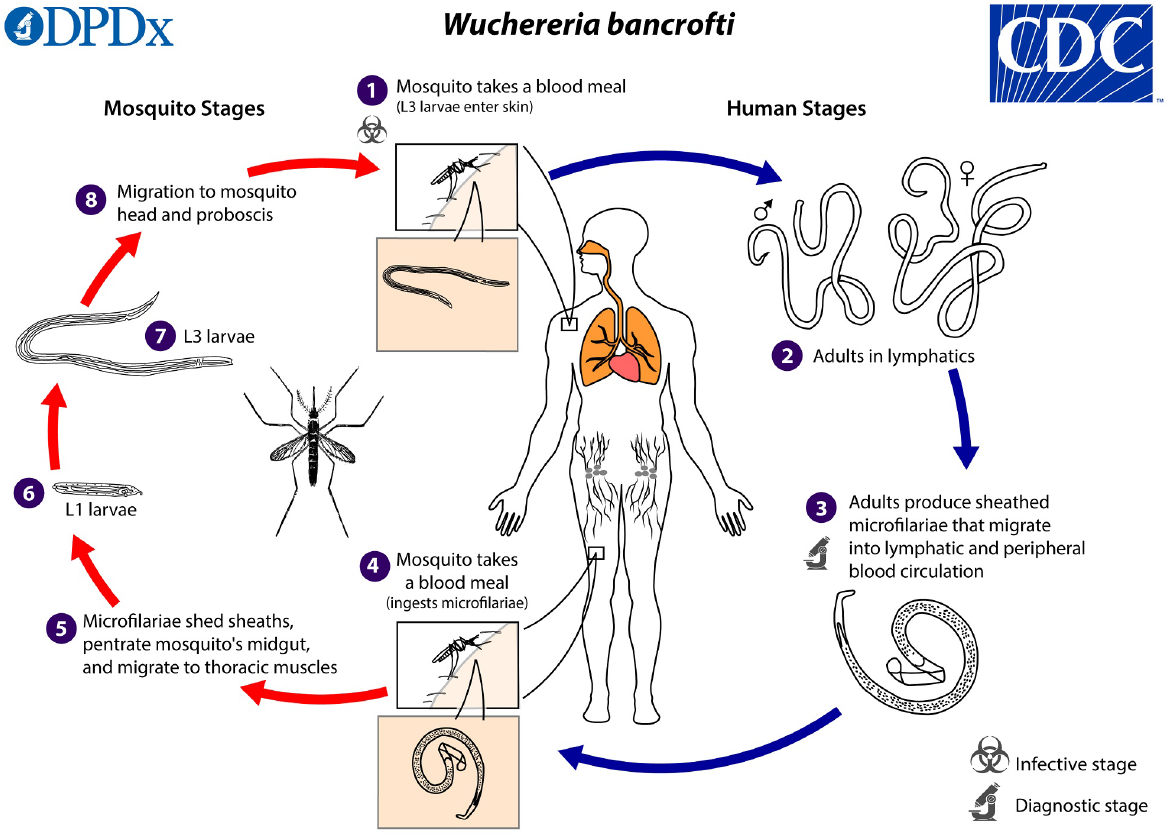
Transmission cycle of the *Wuchereria bancrofti* parasite. Source: Courtesy of Public Health Image Library, Centers for Disease Control and Prevention https://www.cdc.gov/dpdx/lymphaticfilariasis/index.html

The main strategy for controlling LF is mass drug administration (MDA), with the antifilarial drugs diethylcarbamazine (DEC) + albendazole (DA) or ivermectin + DEC + albendazole (IDA) to the entire population at risk of LF, with an exception of pregnant women, children *<*2 years of age, and severely ill patients [8, 12]. MDA with an annual single dose of DEC was implemented in India in 2004 under the National Programme to Eliminate Lymphatic Filariasis (ELF) covering 202 of the 256 endemic districts [13, 14]. Subsequently, DA was introduced in 2007 and all 256 districts were covered by 2008. In 2018, IDA was rolled out in certain districts in phases. To further improve surveillance and programme implementation of MDA, a block-level Implementation Unit (IU) strategy for MDA was adopted in 2022. As of 2023, the programme has made significant progress by interrupting LF transmission through MDA in 139 of the 345 endemic districts, which are currently in the post-MDA transmission assessment surveillance phase. The number of districts under MDA was increased to 345 due to administrative division of the districts and confirmatory mapping of the non-endemic districts. In 2024, of the 1979 IUs in 159 districts where night blood survey was conducted prior to MDA, 57% reported Mf rates below 1%. Efforts to control the mosquito population and reduce human-mosquito contact with the use of bed nets and insecticide-treated materials also contribute to the prevention of LF [11].

Several mathematical models have been proposed to study the transmission dynamics and mitigation strategies of LF [15–17]. The 3 main models which have been widely used to inform public policy for LF are EPIFIL - an age-structured mean worm burden model [18, 19], LYMFASIM - an individual-based stochastic model [20], and TRANSFIL - a stochastic equivalent to the deterministic EPIFIL model that does not account for immunity [21]. Both EPIFIL and LYMFASIM have mechanisms to regulate infection in humans (e.g. immune response) and vectors (density-dependent parasite regulation via parasite induced vector mortality). These 3 models have been used to calculate the required number of MDA rounds to achieve LF elimination under different scenarios and have been instrumental in validating the feasibility of LF elimination as a public health problem through MDA and vector control interventions [19, 21, 22]. WHO recommends 5 rounds of DA-MDA with at least 65% coverage of the total at-risk population [23] to achieve elimination threshold of *<* 1% Mf rate. However, the required number of MDA rounds may vary, depending on the Mf prevalence of a particular endemic region, the effective coverage fraction of MDA [24], and the disease dynamics in that region. For example, Mf prevalence in India has been reported to vary between 2.5% to 10.5% [25–27]. In this context, mathematical models can serve as a useful tool to assess the prospects of LF elimination with antifilarial drugs [28], and have been used to provide an estimate of the number of MDA rounds required to achieve the elimination threshold of *<*1% Mf rate [15].

Several population-level compartmental models have been developed globally to understand the transmission dynamics of infection in humans and mosquitoes [16, 17, 29– 31]. However, most of these models do not consider all the epidemiological phases of LF transmission and have not explored the effects of MDA in detail within such models. The only compartmental model to consider MDA implements a constant MDA effect (i.e., constant proportion of the infected population recovers throughout the year) and highlights the importance of maintaining high MDA coverage per year to reduce the infected population [30]. Further, to the best of our knowledge, none of the individual based or population level models have been applied to estimate the disease burden of LF.

In this paper, we propose a new model - COmpartmental Modelling of Elimination strategies and Transmission of Lymphatic Filariasis (COMET-LF) - with relevant epidemiologic phases and further extend it to incorporate a realistic MDA effect. It is worth noting that most of the population-level models [16, 17, 29, 31] have not considered MDA effects. Consistent with the EPIFIL model, COMET-LF considers a realistic approach to model the effect of MDA, i.e., we consider MDA coverage during a month distributed equally over 4 weeks. Thus, COMET-LF uses a time-dependent MDA effect which is non-zero for 4 weeks and remains zero otherwise. COMET-LF can be used to project the required number of MDA rounds to reduce the Mf prevalence *<* 1% as well as estimate the disease burden (lymphoedema and hydrocele) based on different baseline scenarios of Mf prevalence and MDA coverage.

Control strategies for LF in real-world scenarios are often further complicated by the high heterogeneity in prevalence rates in neighbouring geographical districts or blocks due to migration of infected individuals. Migration can affect the number of MDA rounds required to achieve elimination and resurgence rates in disease-free areas. The COMET-LF model allows us to study these different scenarios. In the final section, we extend our model to a metapopulation model and quantify the risk of LF transmission in a disease-free patch adjacent to an endemic one. Recently, Rajaonarifara et. al. [32] found that migration from surrounding areas may increase the risk of resurgence in Madagascar using LYMFASIM. However, EPIFIL, TRANSFIL and population-level models [16, 17, 29–31] have not been extended to assess the transmission risk in a disease-free area adjacent to an endemic one.

## 2 Methods

### 2.1 Description of COMET-LF

We propose a compartmental model with both human and vector (mosquito) compartments, with births and deaths (Fig. 2). We consider six human compartments, namely, susceptible humans (*S*_*h*_), exposed humans with L3 larvae (*E*_*h*_), those with adult worms (*W*_*h*_), asymptomatic infectious humans with Mf and adult worms (*M*_*h*_), humans with symptomatic disease (*C*_*h*_) and recovered humans (*R*_*h*_). For simplicity, the *C*_*h*_ compartment represents humans with lymphoedema or hydrocele. We adopt this simplifying assumption because the main objective of this article is to evaluate the impact of MDA on the elimination of LF. Therefore, the total human population of the system at any given time *t* is *N*_*h*_(*t*) = *S*_*h*_(*t*) + *E*_*h*_(*t*) + *W*_*h*_(*t*) + *M*_*h*_(*t*) + *C*_*h*_(*t*) + *R*_*h*_(*t*). Susceptible humans become exposed through the bite of an infected female mosquito at a rate *λ*_*vh*_ and move to the *E*_*h*_ compartment. The force of infection from mosquitoes to humans, *λ*_*vh*_, is assumed to be the product of the average mosquito bites per human per unit time 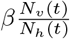, the probability that the mosquito is infected 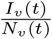, and the probability of successful transmission of infection from vector to human *θ*_*vh*_, i.e. 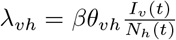. Susceptible humans also increase at a rate Π_*h*_ due to recruitment through new births. Exposed humans may recover at a rate *ω*_*E*_ and move to the *R*_*h*_ compartment. Else, the L3 larvae can grow to become adult worms and thus a human in the *E*_*h*_ compartment may transfer to *W*_*h*_ compartment at rate *λ*_*E*_. From *W*_*h*_, individuals can recover (rate *ω*_*W*_), or transfer to *M*_*h*_ at a rate *λ*_*W*_ if adult worms mate to produce Mf. Both humans with a history of adult worms and Mf can develop symptomatic disease (lymphoedema or hydrocele). We assume humans in the *W*_*h*_ compartment can move to *C*_*h*_ at a rate *α*_1_, while humans in the *M*_*h*_ class move to *C*_*h*_ at a rate *α*_2_. Recovered humans are assumed to have temporary immunity to reinfections. Humans in the *R*_*h*_ compartment lose immunity at a rate *i* and become susceptible again. We assume that all the human compartments decrease due to natural death at rate *μ*. The natural birth and death rates were taken from the Sample Registration System [33] for Bihar.

**Fig 2.**
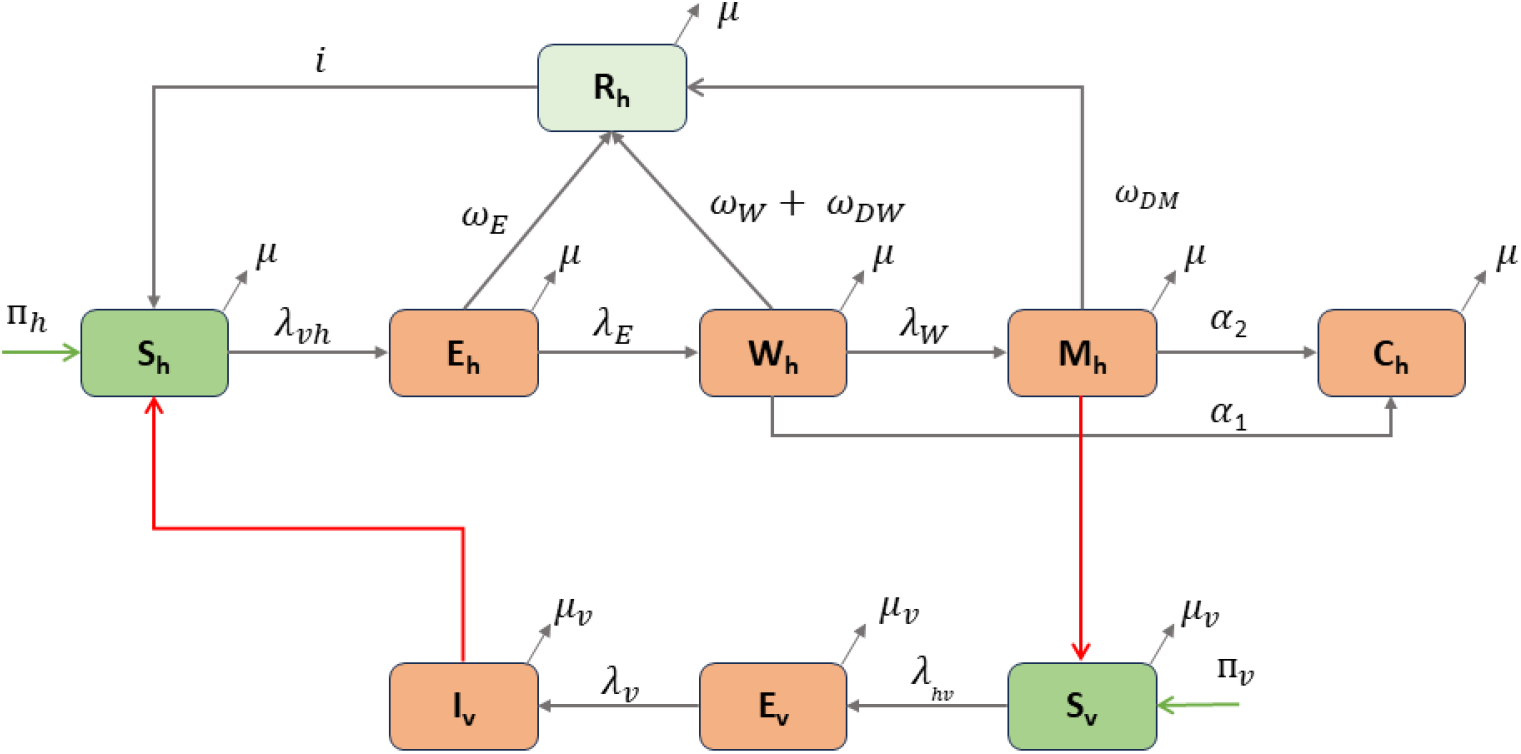
Flow diagram of COMET-LF. Susceptible humans (*S*_*h*_) become infected with L3 larvae (*E*_*h*_) through bites of infectious mosquitoes (*I*_*v*_). They can either self-cure (*R*_*h*_), or become hosts to adult worms (*W*_*h*_). Adult worms can mate to produce microfilaria (*M*_*h*_). Susceptible mosquitoes (*S*_*v*_), which take a blood meal from these humans, become exposed (*E*_*v*_) and eventually become infectious. Humans with a history of adult worms or Mf can develop symptomatic disease (lymphoedema or hydrocele), represented in compartment *C*_*h*_. MDA effects are modeled through additional recovery rates *ω*_*DW*_ and *ω*_*DM*_ of the *W*_*h*_ and *M*_*h*_ compartments. In the absence of MDA, these two rates are set to zero.

The vector population is divided into 3 epidemiological compartments: susceptible vectors (*S*_*v*_), exposed vectors with Mf (*E*_*v*_), and infectious vectors (*I*_*v*_) (Fig. 2). Therefore, the total vector population at any given time *t* is *N*_*v*_(*t*) = *S*_*v*_(*t*)+*E*_*v*_(*t*)+*I*_*v*_(*t*). Susceptible mosquitoes become exposed when engorging on the blood of infectious humans at a rate *λ*_*hv*_. The force of infection from humans to mosquitoes 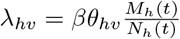, which is the product of the average number of bites per mosquito per unit time *β*, the probability that the human is infectious 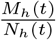, and the probability of successful transmission of infection from human to vector *θ*_*hv*_. Additionally, susceptible mosquitoes are recruited at a rate Π_*v*_ due to new births. Exposed mosquitoes may become infected at a rate *λ*_*v*_. We assume that all mosquito classes decrease due to natural death at a rate *μ*_*v*_.

The mathematical equations governing the dynamics of infection are detailed in S1A text. The description of model parameters and their corresponding estimates from published literature or expert opinions are reported in Table 1.

**Table 1.** Fixed model parameter descriptions and their estimates from literature.

| Parameter | Description | value/Range | unit | Reference |
| --- | --- | --- | --- | --- |
| $N_h(0)$ | Initial human population | 100000 | humans | Assumed |
| $N_v(0)$ | Initial mosquito population | $n_v N_h(0)$ | Mosquitoes | [17] |
| $n_v$ | Ratio of mosquito population and human population | 3 | unitless | [17] |
| $\Pi_h$ | Recruitment rate of humans | $N_h(0) \times b_h$ | human week<sup>-1</sup> | |
| $\mu$ | Natural death rate of humans | 0.0067/52 | week<sup>-1</sup> | [33] |
| $b_h$ | Birth rate of humans | 0.0277/52 | week<sup>-1</sup> | [33] |
| $\Pi_v$ | Recruitment rate of mosquitoes | $N_v(0) \times b_v$ | mosquito week<sup>-1</sup> | – |
| $\mu_v$ | Natural death rate of mosquitoes | 1.25 | week<sup>-1</sup> | [19] |
| $b_v$ | Birth rate of mosquitoes | 1.25 | week<sup>-1</sup> | Assumed<br>= $\mu_v$ |
| $\theta_{hv}$ | Transmission probability from humans to vectors | 0.37 | unitless | [19] |
| $\beta$ | Average bite per mosquito per week | 2.5 | week<sup>-1</sup> | [19] |
| $\lambda_v$ | Progression rate of latent mosquitoes to infected mosquitoes | 7/11 | week<sup>-1</sup> | [9] |
| $i$ | Immunity waning rate | 0.000004 | week<sup>-1</sup> | [37] |

For the COMET-LF model, we derive the expression of the unique disease-free equilibrium (DFE) of the model. Using the next-generation matrix method [34], we calculate the basic reproduction number (*R*_0_) as follows:

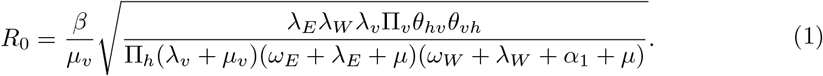

Using dynamical system theory, we prove that the DFE of the model (S1) is globally asymptotically stable whenever *R*_0_ *<* 1. We also calculate the endemic equilibria (EE) and their local stability conditions. The proofs of these analytical results are given in S1B and S1C text.

An important intervention in the context of LF elimination is MDA. We investigate the effect of MDA in our model for different baseline scenarios of Mf prevalence. MDA campaigns are conducted annually, and we incorporate this through a time-dependent impulsive MDA function *γ*(*t*). If 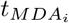 is the first week of conducting *i*^*th*^ MDA, then the impulsive MDA function is given by,

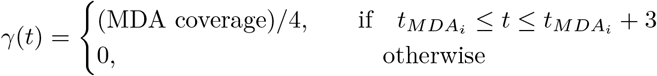

This approach is consistent with existing EPIFIL and LYMFASIM models, which also consider MDA activities conducted in a particular month of a given year [35]. Infectious humans in the *M*_*h*_ compartment are assumed to move to the *R*_*h*_ compartment at a rate *ω*_*DM*_ due to MDA. Similarly, humans in the *W*_*h*_ compartment (adult worms but no Mf) move to *R*_*h*_ at a rate *ω*_*DW*_ due to MDA. We assume *ω*_*DM*_ = *p*_1_*γ*(*t*), and *ω*_*DW*_ = *p*_2_*γ*(*t*), where *p*_1_ models the effect of the drug on clearing Mf and adult worms (proportion of persons cured from Mf and adult worms, cure rate), while *p*_2_ is the effect of the drug on adult worms (proportion of persons that cleared adult worms). We consider both the DA and IDA drug regimens for simulation. We use human clinical trial data in the Indian setting to model the effects of MDA in the COMET-LF model. For DA and IDA, the values for *p*_1_ are taken as 61.8% and 84%, respectively [14], while *p*_2_ is taken as 15% [14] for both the drug regimens. This approach is in contrast to EPIFIL and LYMFASIM which models MDA effects by considering the percentage of Mf clearance, percentage of worms killed and worm sterilisation periods as reported in Irvine et. al., [35]. The full details of the MDA implementation in COMET-LF are provided in S1D text.

### 2.2 Model quantification and scenario analysis

For the calibration of the disease progression parameters, *α*_1_ and *α*_2_, we calibrate time series data using the MATLAB function ‘*fminsearchbnd*’ to minimise the sum of squared error between baseline and model generated outputs of Mf rate and disease rate. *α*_1_ and *α*_2_ are allowed to vary in the interval (0, 1). These estimates of *α*_1_ and *α*_2_ remain unchanged throughout the rest of the paper. Table 1 shows the list of known parameters for which estimates are taken from the literature.

We calibrate the model in the absence of MDA (*ω*_*DM*_ = *ω*_*DW*_ = 0) using publicly available annual Mf rates and symptomatic disease rates for Bihar for the period 2008 – 2013 [36]. In the calibration step, we estimate seven parameters – the probability of successful transmission of infection from vector to human *θ*_*vh*_, the progression rate from humans with L3 larvae to humans with adult worms *λ*_*E*_, the self-cure rate of exposed humans in absence of MDA *ω*_*E*_, the rate at which humans with adult worms develop Mf *λ*_*W*_, the self-cure rate of humans with adult worms in absence of MDA *ω*_*W*_, and the two disease progression rates *α*_1_ and *α*_2_ from the *W*_*h*_ and *M*_*h*_ compartments, respectively. All the rates are allowed to vary in the interval (0, 1), while *θ*_*vh*_ lies in the range (0, 0.37) [19].

For the scenario analysis, the calibrated model was applied to simulate different MDA-scenarios (2 drug regimens, 5 levels of MDA coverage: 45%, 55%, 65%, 75% and 85%) for four different endemic scenarios (1.5%, 2%, 5% and 10%). Thus, 10 MDA scenarios were simulated for each endemic scenario. The parameters are the probability of successful transmission of infection from vector to human *θ*_*vh*_, the progression rate from humans with L3 larvae to humans with adult worms *λ*_*E*_, the self-cure rate of exposed humans in the absence of MDA *ω*_*E*_, the rate at which humans with adult worms develop Mf *λ*_*W*_, the self-cure rate of humans with adult worms in absence of MDA *ω*_*W*_, and the two disease progression rates *α*_1_ and *α*_2_ from the *W*_*h*_ and *M*_*h*_ compartments, respectively. All the rates were allowed to vary in the interval (0, 1), while *θ*_*vh*_ *ϵ* (0, 0.37) [19]. We produced 100,000 initial samples using Latin Hypercube Sampling. For each baseline scenario, we identified 250 samples that fall within the required Mf rate with an allowable error of *±*0.5% (e.g. 1% - 2% for the 1.5% baseline). We identified samples of parameters that lead to the baseline Mf rates 1.5% (1% - 2%), 2% (1.5% - 2.5%), 5% (4.5% - 5.5%) and 10% (9.5% - 10.5%). The identified samples were then used to generate future trends of Mf rates using the model. The number of MDA rounds required to achieve *<* 1% Mf rate with a 99% probability was obtained. The probability of elimination is calculated by dividing the number of trajectories *<* 1% Mf rate after 1 year of stopping MDA by the number of all trajectories for each simulation scenario.

### 2.3 Impact of MDA on disease (lymphoedema and hydrocele) prevalence

Using the previously obtained equilibrium fits to different levels of baseline Mf rates, 10% and 2% (Sec 2.2), we simulated 6 scenarios to assess the impact of 5 annual rounds of MDA (DA or IDA) for 3 different MDA coverages (45%, 65% and 85%). We predicted the trends in disease prevalence over time before and after stopping MDA over a period of 20 years.

### 2.4 Implementation of EPIFIL and LYMFASIM

LYMFASIM [20] is based on the stochastic microsimulation of individuals. This model simulates life histories of humans including birth, death and acquisition/loss of parasites. Additionally, the individual parasites are also simulated including their maturation, mating, Mf production, and death. The model simulates population changes by replicating demographic processes. Births occur randomly based on age-specific fertility rates and the number of females in each age group. Deaths occur randomly as per age-specific life tables for India. Each individual in the simulation has fixed traits such as gender and susceptibility to infection, determined by random selection from preset probability distributions. Other traits, such as fertility rates and exposure to mosquitoes, can change during the simulation. The model focuses on simulating parasite transmission and monitors changes in infection status (e.g. number of immature/mature; male/female worms) in individuals over time. The output comprises results of simulated epidemiological surveys to be conducted at user-defined moments (year and month). The following outputs can be generated: i) population-level summary; ii) population-level details by age and sex; and iii) individual-level metrics. From these, we can calculate the required number of MDA rounds to reach the elimination threshold (*<* 1% Mf rate) in specific scenarios. At present, the model does not account for immigration or emigration of humans.

EPIFIL [18] is a deterministic model that simulates the dynamics of LF. The model comprises a system of partial differential equations coupled with a single ordinary differential equation describing the patterns of infection over age and time. Three state variables - the mean worm burden, the mean Mf intensity, and the mean acquired immunity level constitute the system of partial differential equations. Mean intensity of infective L3 larvae per mosquito is described by an ordinary differential equation. This model is also capable of calculating the required number of MDA rounds to reach the elimination threshold.

We applied both EPIFIL and LYMFASIM to simulate the impact of a single dose of DA and IDA, administered annually, for different MDA-scenarios considered in COMETLF. For both models, we used the respective model estimates for all the biological parameters, representing Indian settings, as described elsewhere [19, 38]. While in LYMFASIM we allowed the mosquito biting rate to vary from 1600 to 3200 per month to represent different endemic scenarios in India, in EPIFIL, we followed the same methodology as COMET-LF to simulate the different endemic scenarios.

In both LYMFASIM and EPIFIL, the efficacy of the drug regimens, DA and IDA, are distinguished by 3 different parameters - worm killing rate, worm sterilisation rate, and Mf clearance rate (Table 2). For both these models, drug efficacies were obtained from Irvine et al [35]. For each scenario: baseline Mf rate (1.5%, 2%, 5% and 10%), MDA-coverage (45%, 55%, 65%, 75% and 85%), and for a fixed round of either DA or IDA, 1000 simulations were conducted. The number of simulations in which an Mf rate was found *<* 1% was considered as the probability of elimination for the scenario. We predicted the number of rounds of MDA required to achieve *<* 1% Mf rate. In this way, we estimated the number of rounds of MDA required for each scenario that would achieve *<* 1% Mf rate with a 99% probability.

**Table 2.** Two drug regimens and their corresponding effects on Mf clearance rate, worm killing rate, and worm sterilisation rate. These values were used to simulate MDA effects from LYMFASIM and EPIFIL models.

| Drug regimen | Mf clearance (%) | Worm killing (%) | Worm sterilisation period (%) |
| --- | --- | --- | --- |
| DA | 95% | 55% | 0% |
| IDA | 100% | 55% | 100% |

### 2.5 Metapopulation model for migration

Metapopulation models have been used in the literature to investigate the effects of migration [39]. In this approach, the population is partitioned into a number of discrete patches or regions. In each region, the disease dynamics follows a compartmental model corresponding to the infection and disease prevalence in this region. Humans can migrate between the distinct regions, leading to a multi-patch multi-compartment model of disease transmission. For our metapopulation model (model description in SI), we consider a two-patch system to investigate the role of migration. We assume that the two patches are connected through migration of susceptible humans, exposed humans, people with adult worms, people with both adult worms and Mf, and recovered people. We introduce a migration rate parameter (*m*) for all the classes of humans. For simplicity, we assume the migration rates of all the classes are the same. A schematic of this model is shown in Fig. 3.

**Fig 3.**
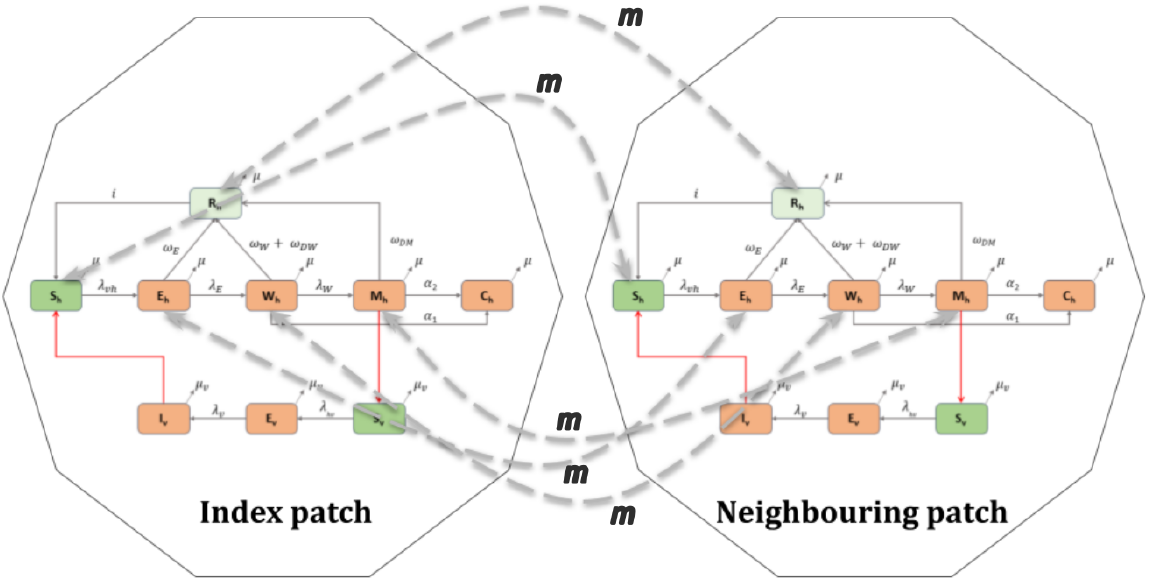
Schematic representation of the two-patch system. Humans in all compartments, except the disease compartment, migrate between the two patches with rate *m*.

The migration rate *m* is calculated based on 2011 census (D-02 series [33]) data for migration within and across states in India. The in- and out-migration rate of Bihar is estimated using the current, last place of residence and all duration of residence data [33]. The in-migration and out-migration rates per 1000 population per 10 years are calculated as:

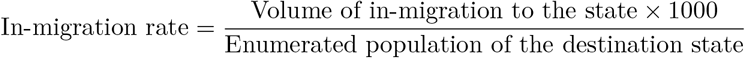

And

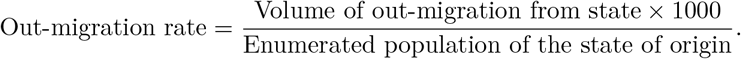

For Bihar, the value of in-migration and out-migration are 11 per 1000 and 72 per 1000, respectively. We use the mean value of these to obtain the migration rate of 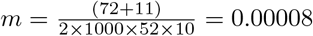 per week for Bihar. To account for demographic changes and temporary migration for seasonal work, we also simulate the model for 2 higher migration rates, *m* = 0.0005 and *m* = 0.001. All other parameters are taken from Table 1 and MDA related parameter values are taken from Section 2.1.

For the simulations that compute additional number of MDA rounds, we consider that the index (or target) location is connected to a neighbouring region, each with its own baseline Mf rate ∈ {1.5%, 2%, 5%, 10%}. The value of the migration parameter is assumed to be 0.001. We apply MDA to both regions with a coverage of 45%. Among the 250 × 250 parameter values and initial conditions, we simulate the metapopulation model with 100 × 100 parameter values and initial conditions. Based on the number of MDA rounds predicted for the given baseline Mf rate in the absence of migration in the neighbouring patch, we calculate the required number of MDA rounds to achieve elimination in the target location. For the simulations, which investigate the risk of resurgence in a non-endemic region, we consider the target location to be an infection-free patch where all humans and vectors are susceptible and couple it to a neighbouring location with a baseline Mf rate of 1.5%, 2%, 5% or 10%. We assume that all disease-related parameters and the total population are the same in both patches. We consider 3 different migration rates, 0.001, 0.0005 and 0.00008, for all the trajectories. We then calculate the risk of Mf transmission in the non-endemic patch by dividing the number of trajectories *>* 1% Mf rate after 5 years in the disease-free block by the number of all trajectories.

## 3 Results

### 3.1 Model calibration and disease (lymphoedema and hydrocele) rates

We calibrate the model in the absence of MDA (*ω*_*DM*_ = *ω*_*DW*_ = 0) using publicly available annual Mf rates and symptomatic disease rates for Bihar for the period 2008 – 2013 [36]. The estimated parameters are *θ*_*vh*_ = 0.3104, *λ*_*E*_ = 0.3945, *ω*_*E*_ = 0.9225, *λ*_*W*_ = 0.0389, *ω*_*W*_ = 0.8087, *α*_1_ = 1.564 × 10^−4^ and *α*_2_ = 4.938 × 10^−5^. The model fits are depicted in Fig. 4(a). We fixed the values of *α*_1_ and *α*_2_ for the rest of the manuscript. For scenario analysis, the calibrated estimates of *α*_1_ and *α*_2_ were used to simulate different endemic scenarios, by calibrating the parameters *θ*_*vh*_, *λ*_*E*_, *ω*_*E*_, *λ*_*W*_, and *ω*_*W*_ to obtain the baseline Mf rate. An example of an equilibrium fit following this procedure for a 5% baseline prevalence is shown in Fig. 4(b). Corresponding to this equilibrium fit, for the estimated values of *α*_1_ and *α*_2_, we obtain an equilibrium for the disease prevalence as well. This is shown in Fig. 4(c) for the baseline Mf of 5%, which yields a disease prevalence in the range 1.75% − 2%. On administration of MDA, Mf prevalence reduces systematically, until it falls below the elimination threshold. An example of the Mf time series is shown for 5% baseline Mf rate and 45% DA-MDA coverage in Fig. 4(d).

**Fig 4.**
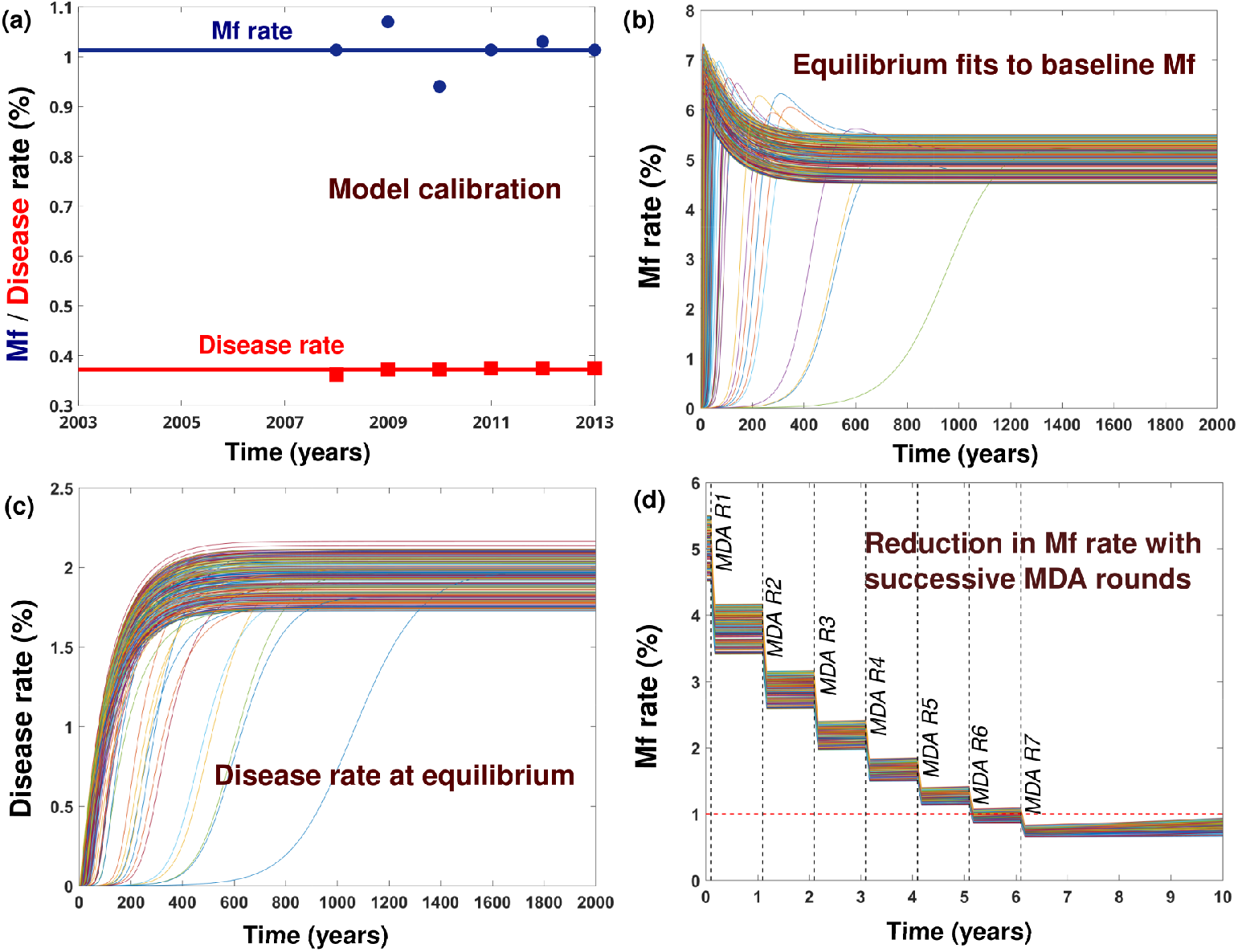
Methodology for projecting the required number of MDA rounds to reach *<* 1% Mf rate. The baseline Mf rate is considered to be 5% (4.5% - 5.5%). (a) Simultaneous fitting of Mf rate and disease rate. (b) Model generated trajectories of Mf rates achieving equilibrium Mf rates between 4.5% - 5.5%. (c) Model generated trajectories of disease rate corresponding to equilibrium Mf rate in previous step. (d) Starting from the equilibrium achieved in previous step, drug regimen DA with 45% MDA coverage is considered to generate future trends of Mf rates. Seven DA-MDA rounds are required to attain the elimination threshold of 1% Mf rate. The dotted red line indicates 1% Mf rate.

### 3.2 Number of MDA rounds required for elimination

The number of rounds of MDA (DA or IDA) required to achieve LF elimination (*<* 1% Mf prevalence) was estimated using all 3 models. The predicted number of rounds with DA or IDA are shown in Fig. 5(a)-(f). The predictions of the COMET-LF model are shown in Fig. 5(a)-(b). As expected, the coverage fraction plays an important role along with the baseline Mf rate in determining the required number of MDA rounds. This effect is especially pronounced if the baseline Mf rate is high, for example, for the DA regimen, at a baseline prevalence of 10%, 5 rounds of MDA are required at 85% coverage, but this number can increase to 10 rounds if the coverage falls to 45%. The corresponding predictions for the IDA regimen at 10% prevalence goes from 4 rounds at 85% coverage, increasing to 7 rounds at 45% coverage. In general, at 85% coverage with DA and IDA, 5 rounds of MDA are sufficient to achieve elimination for baseline Mf prevalence up to 10%.

**Fig 5.**
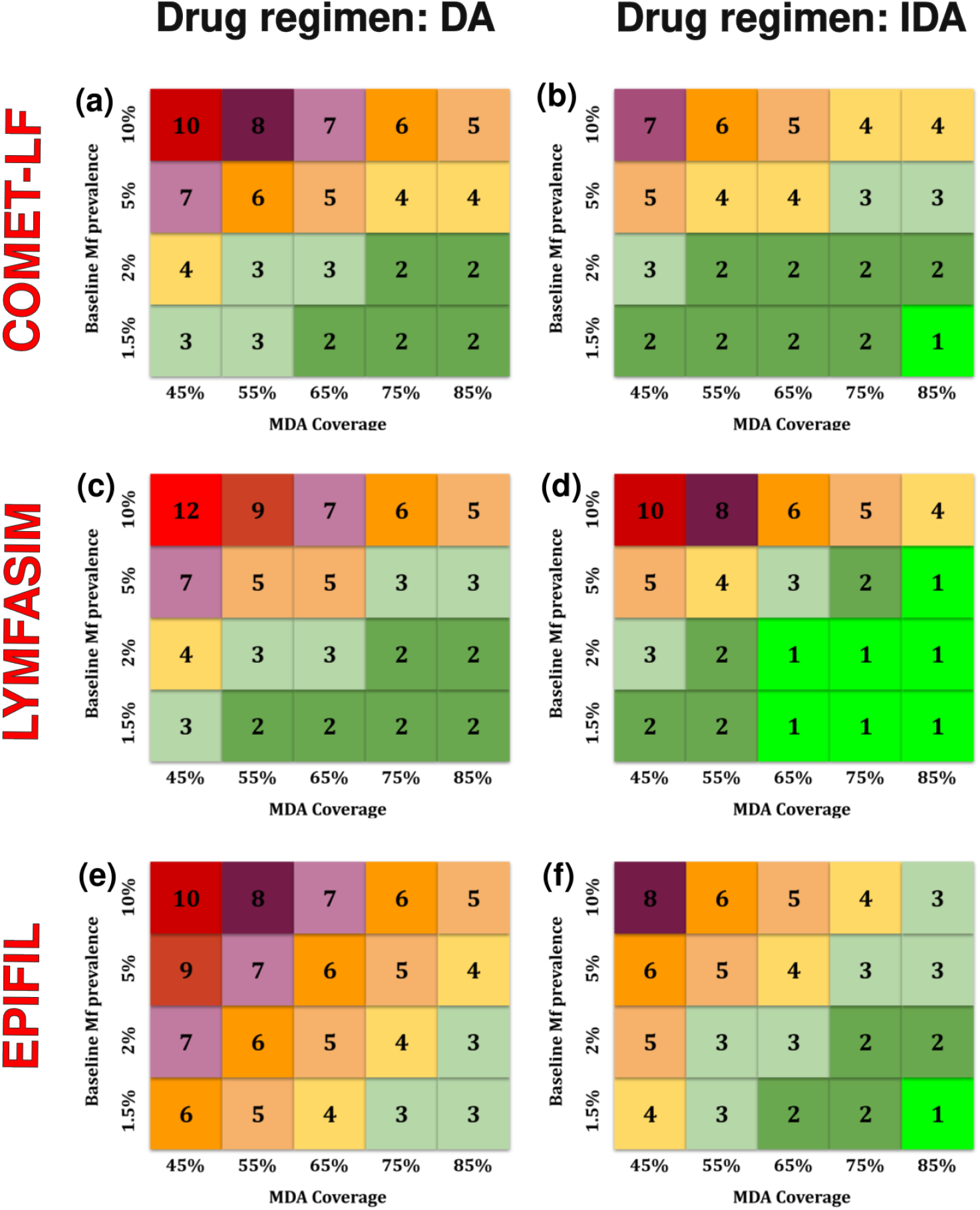
Predicted number of MDA rounds required to achieve 99% probability of elimination with DA (left panel) and IDA regimens (right panel) by COMET-LF (a)-(b), LYMFASIM (c)-(d) and EPIFIL (e)-(f).

In order to validate the predictions of the COMET-LF model, we compared these projections to those obtained from EPIFIL and LYMFASIM models for DA and IDA. WHO recommends 5 rounds of annual MDA using DA with 65% MDA coverage of the total at-risk population to achieve elimination [23]. We observed this consistently in all 3 models studied (Fig. 5). For DA, the predicted number of rounds were within ±1 range for COMET-LF and LYMFASIM, with the only deviation occurring for very high prevalence (Mf = 10%) with very low coverage (45%), where LYMFASIM predicts two extra rounds. In contrast, while EPIFIL prediction were similar for high Mf rates, there were significant deviations compared with COMET-LF and LYMFASIM predictions (Fig. 5) for low Mf rates (±3 rounds). On the other hand, for the 3-drug IDA protocol, predictions for all 3 models were between ±2 rounds. For high coverage and moderate-to-low Mf rates, COMET-LF predicts 1-2 additional rounds of MDA compared to LYMFASIM, while for low coverage and very high Mf rates, LYMFASIM predicts 2-3 additional rounds (Fig. 5). EPIFIL, in contrast, predicts 1-2 additional rounds compared to COMET-LF. For all 3 models, MDA with DA requires 1-3 additional rounds compared to IDA for most of the scenarios. The probability of achieving the elimination threshold as a function of the number of rounds of MDA is shown for each drug regimen and endemic scenario in Figs. S1 - S2.

### 3.3 COMET-LF for assessing MDA impact on disease (lymphoedema and hydrocele) prevalence

The COMET-LF model allows us to generate estimates of disease (lymphoedema and hydrocele) prevalence consistent with different equilibrium baseline Mf rates. The values of the disease progression rates from infectious humans are fixed at *α*_1_ = 1.564 × 10^−4^ and *α*_2_ = 4.938 × 10^−5^ from the model calibration. We simulated four different scenarios with baseline Mf rates at 1.5%, 2%, 5% and 10%. The disease prevalence (lymphoedema and hydrocele) ranged from 0.57% ± 0.3 % at a baseline Mf rate of 1.5 % to 3.83 % ± 0.2 % at a baseline Mf rate of 10%.

There is a systematic decrease in the disease prevalence on administration of 5 rounds of MDA. The distributions of disease rate with respect to the number of years after starting MDA is shown in Fig. 6 and in Fig. S3. First, we note that while elimination of Mf happens over a 5-10 year timescale, even for high baseline Mf rates, the disease dynamics persist for much longer timescales. This is independent of the MDA coverage and the drug regimen. The left (right) hand column of Fig. 6 shows the reduction in disease prevalence over a 20 year timescale for the DA and IDA regimens with 65% and 85% coverage, and 10% (2%) baseline Mf rate. While increased coverage and shift to IDA regimen offers marginal improvements, the long timescales involved imply these gains are only incremental. For the 2% baseline Mf rate, since the disease rates are initially low, the introduction of MDA results in only a slow decline. Relative reductions in disease prevalence are roughly equal between different baseline Mf rates, with a reduction of ≈ 10.5% for DA and ≈ 11.5% for IDA at 85% coverage at 20 years following the start of MDA. For a low MDA coverage of 45%, the corresponding reduction was ≈ 8% for DA and 9.5% for IDA (Fig. S3).

**Fig 6.**
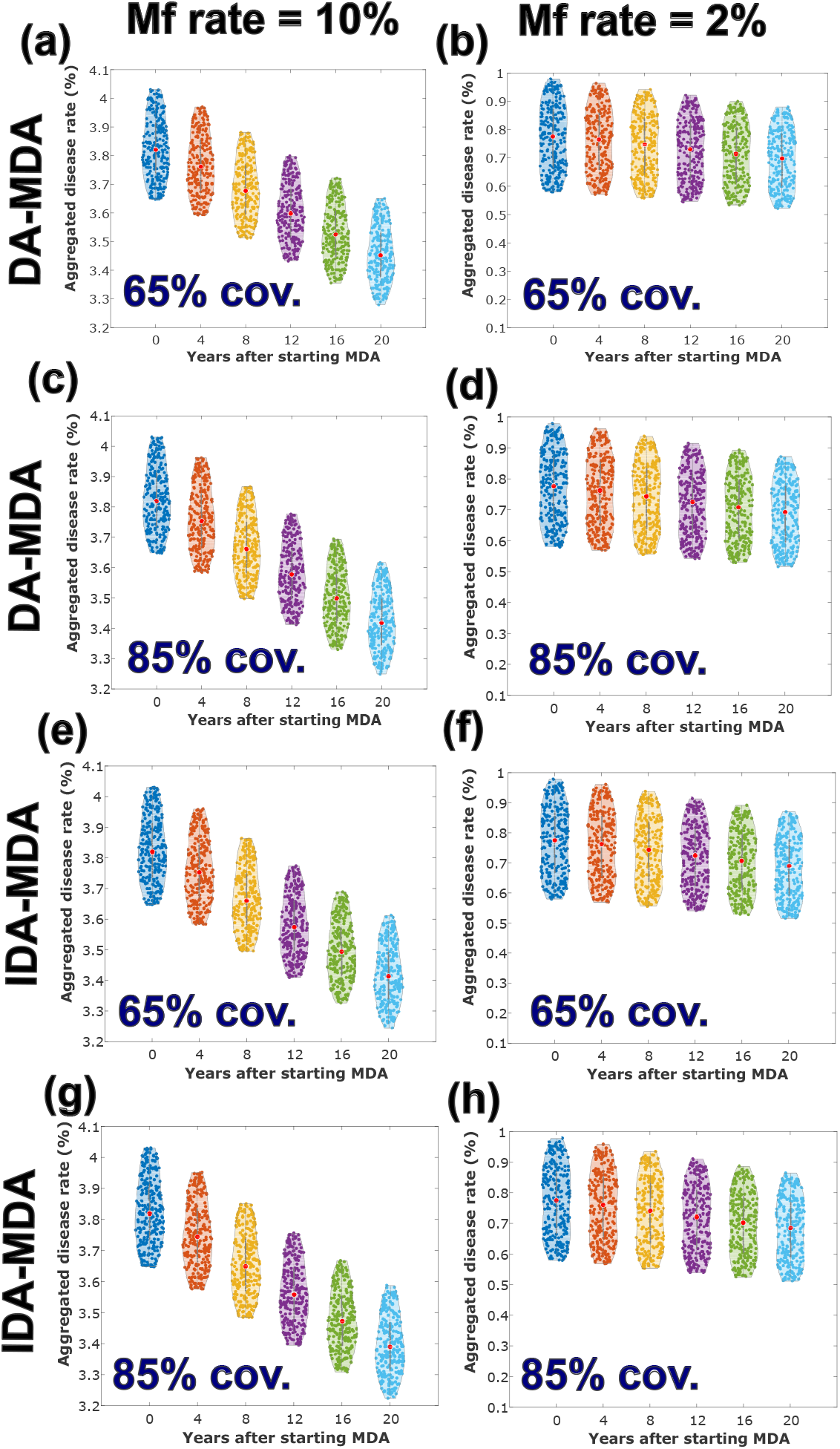
Projection of disease prevalence corresponding to two different initial baseline Mf rates of 10% (left column) and 2% (right column). MDA was applied for 5 years according to the following protocols, DA with 65% coverage (a-b), DA with 85% coverage (c-d), IDA with 65% coverage (e-f), and IDA with 85% coverage (g-h). In each violin plot, the red circle in the middle represents the median value of the distribution whereas the grey bars spans from first to third quartile (interquartile range). The thin grey lines at the boundary represent the rest of the distribution.

### 3.4 The role of migration in LF elimination

A major challenge in the elimination of communicable diseases is the migration of human hosts to neighbouring spatial units and its impact on transmission of infection [40]. This is of particular importance in the case of diseases such as LF, where the asymptomatic infectious phase (humans with Mf) may last for 10 years or longer [41], thus allowing for transmission. We investigated a two-patch system to study different scenarios. In each region, the disease dynamics follows a compartmental model corresponding to the infection and disease prevalence in the region. Humans can now migrate between the distinct regions, leading to a multi-patch multi-compartment model of disease transmission.

We first consider how the predicted number of MDA rounds required for elimination can change if the target location neighbours another endemic region with a different Mf rate. We then calculate the number of MDA rounds required to achieve elimination in the index region in the presence of migration and compute the difference from the prediction in Fig. 5. We do this for both DA and IDA drug regimens. For the DA drug regimen, as shown in Fig. 7(a), our results suggest that the predicted number of MDA rounds can increase by up to 2-3 additional rounds depending on the Mf prevalence in the neighbouring region. For example, when migration was not considered, 3 rounds of DA-MDA were predicted to be required for elimination at a baseline Mf rate of 1.5%. However, if the spatial region neighbours a high prevalence region with a baseline Mf rate of 10%, the required number of DA-MDA rounds increases to 5. Although the IDA drug regimen fares better, even in this case there can be an additional requirement of 1-2 rounds of MDA, particularly if the Mf rate in the neighbouring region is very different, as shown in Fig. 7(b).

**Fig 7.**
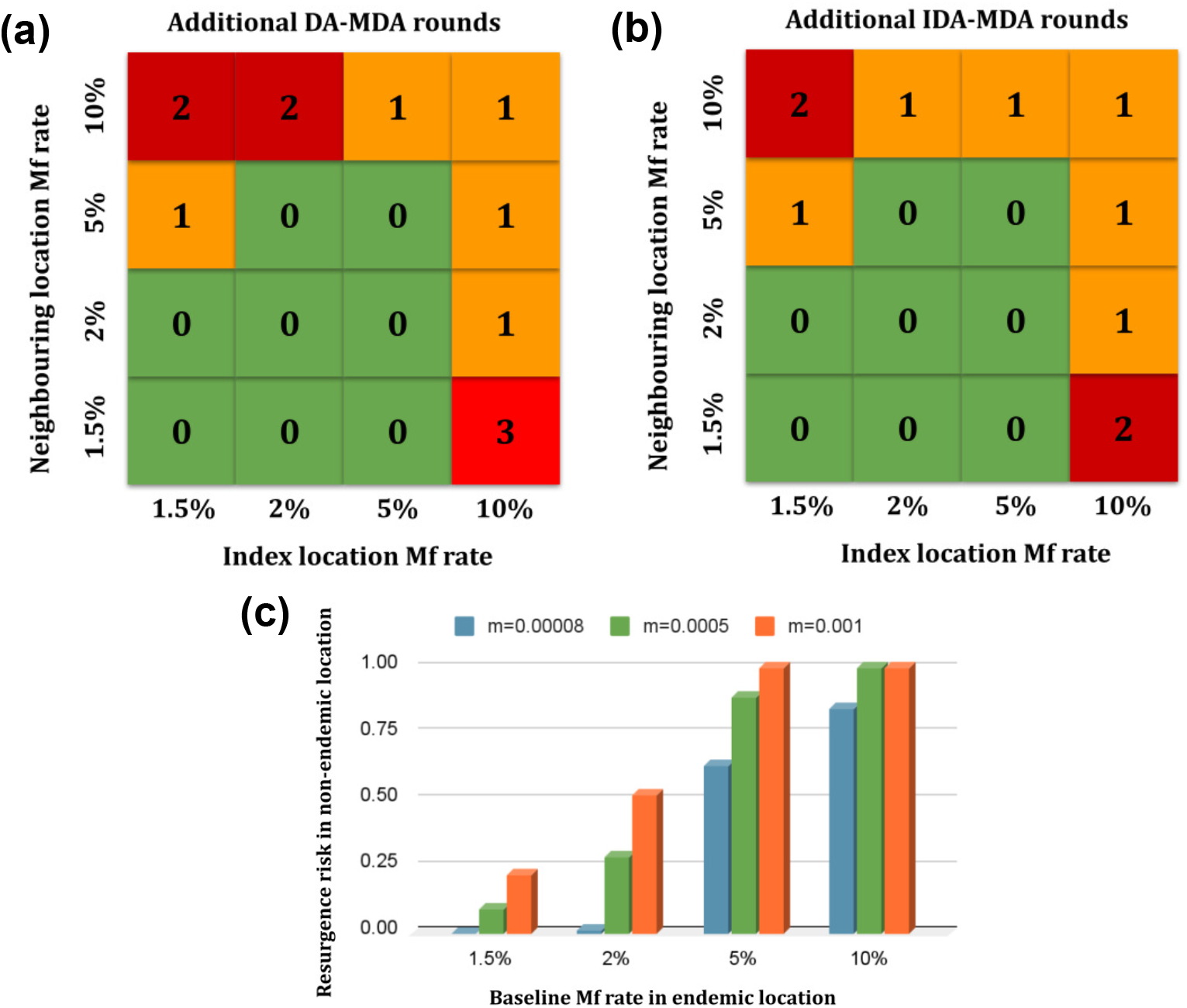
(a) Additional rounds of DA-MDA required in an index location adjacent to a neighbouring endemic region at a coverage of 45%. (b) Additional rounds of IDA-MDA required in an index location adjacent to a neighbouring endemic region at a coverage of 45%. (c) Risk of Mf transmission to a non-endemic patch after 5 years given the endemic patch has reached a baseline Mf rate for 3 migration rates *m* = 0.00008, *m* = 0.0005, and *m* = 0.001.

Migration can also impact resurgence or a shift to the EE in previously non-endemic areas. To understand this effect, we considered a non-endemic region which neighbours a region with endemic LF infection. We study this for 3 different migration rates of 0.00008, 0.0005 and 0.001 to determine the probability that the previously non-endemic region becomes endemic (Mf rate *>* 1%) within a period of 5 years, as shown in Fig. 7(c). If the neighbouring location has a very low Mf rate, the probability of resurgence in the index patch remains low, with *≈* 22% probability for a high migration rate *m* = 0.001. Even at an Mf rate of 5%, the resurgence probability increases to 63% for a low migration rate (*m* = 0.00008) and to 100% for high migration rates. Alarmingly, if the Mf prevalence in the neighbouring patch is high (*>* 5%), resurgence probability rises above 75% within a 5-year timescale for all 3 migration scenarios.

## 4 Discussion

We propose a new population-level compartmental model - COMET-LF - for the transmission of LF that accounts for humans with L3 larvae, those with adult worms, and infectious humans with Mf. Additionally, the model captures humans with symptomatic disease. The dynamical properties of the model were analysed to determine DFE and the EE. *R*_0_ was computed and the stability of the DFE was assessed (S1B).

COMET-LF was used to predict the required number of MDA rounds to achieve *<*1% Mf rate for elimination of LF, using both DA and IDA drug regimens and different levels of drug coverages. The predictions were compared to the established mean worm-burden model EPIFIL [19] and the stochastic miscrosimulation model LYMFASIM [20]. COMET-LF considers the population-level epidemiology of LF within a deterministic framework, which is distinct from EPIFIL and LYMFASIM. The consistency of the predictions with the aforementioned established models provide credence to the COMETLF model (Fig. 5). The model can be used to make tailored predictions of the required number of MDA rounds for various endemic regions in the country.

In contrast to existing models of LF transmission [17, 18, 20, 21, 30, 31], COMET-LF can generate estimates of disease prevalence given the baseline Mf rates were achieved at equilibrium. The disease burden increases with increasing baseline Mf rate. However, more specific calibrations are required to assess the actual burden in a community. Next, we investigated the effect of 5 MDA rounds on the disease rate (Fig. 6 and SI Fig. S3). We observe that MDA coverage plays a crucial role in reducing the disease prevalence in a community. For instance, 5 rounds of DA-MDA with 85% coverage exhibits around 10.5% reduction in mean disease rate within 20 years of starting MDA.These results are comparable with a previous Indian study where a 14% reduction in hydrocele was reported after 7 rounds of DEC alone [42]. However, in that study, the decline of lymphoedema was minimal. Lymphoedema develops gradually over a long period, and reducing the disease burden relies on early case identification, management, and treatment. Hydrocele burden can be reduced if all cases undergo hydrocelectomy. MDA coverage alone is insufficient to lower the disease burden; treatment compliance, including proper consumption of MDA drugs during the campaign and ensuring all previously untreated cases are addressed, is essential. More accurate estimates can be generated in the future by assessing both the Mf and disease prevalence in specific areas to calibrate the model to generate localised predictions.

A major concern for the elimination of communicable diseases is the spatial heterogeneity and the transmission of infection from areas of high endemicity to neighbouring regions [40]. This is of practical interest since endemic blocks or districts are often clustered together in close spatial proximity in many regions of India, and effective coverage can often be low. We report for the first time, within a meta-population two-patch extension of the COMET-LF model, the putative effect of migration on the number of MDA rounds required for elimination. For the DA drug regimen, the required number of rounds can increase by upto 2-3 additional rounds, while for the IDA regimen these can increase by 2 additional rounds, depending on the baseline prevalence and MDA coverage. These deviations from the recommended number of rounds may explain the relative slow decline in Mf rates in India, where MDA programs, especially using the two-drug DA regimen have been operational since 2007 [13]. Furthermore, we show that if a disease-free patch is spatially adjacent to an endemic region, migration allows for the resurgence of Mf in the previously disease free region with a high probability in different baseline scenarios (Fig. 7). Again, this is of practical concern when a region where LF has been eliminated lies within a spatial cluster of high endemicity. Although this scenario analysis assumes the two patches to be identical demographically, and in reality, various location-specific factors can affect transmission, our work points to the crucial role migration can play in achieving elimination. Future work, incorporating more realistic demographic characterisation of specific locations can then be used to assess the impact of migration in targeted areas.

As a deterministic population-level compartmental model, COMET-LF has some limitations, which can be addressed in future extensions of the model. The model assumes that each infected human has the same number of adult worms and Mf. In reality, the long duration of the asymptomatic phase of LF implies that there is considerable heterogeneity in the worm and Mf burden, and this can impact the transmission dynamics as well as the effect of MDA. Furthermore, there is considerable worm heterogeneity across various age groups, which has not been captured in the model [24]. Additionally, we have not accounted for parasite-induced vector mortality in the model [43, 44]. Although we generate aggregated disease burden, disaggregated lymphoedema and hydrocele compartments will be able to provide more accurate burden estimates. In addition, hydrocelectomy also plays a significant role in the reduction of disease prevalence, which has not been taken into account. In future work, the model can be extended to account for socio-economic characteristics, change in treatment regimens, increase in MDA coverage over time, data quality issues, and the role of health system strengthening efforts. Finally, we assume a homogeneous MDA coverage; in reality, there can be groups of individuals who do not participate in MDA. The never-treated group can pose a significant challenge in achieving the elimination target. In the absence of any systematic review on the efficacy of DA and IDA, we use data from a clinical trial that was conducted in India [14].

In summary, we present a simple compartmental model that can capture the transmission dynamics of LF and can project the required number of MDA rounds to reach elimination. Our results point to the critical role of MDA coverage in achieving elimination. COMET-LF can also provide estimates of disease burden in a community. The two-patch COMET-LF model results are also helpful in designing more accurate control strategies. For instance, the migration from an endemic region can be screened for Mf to reduce further transmission to non-endemic regions. Importantly, the COMET model has the flexibility to be applied to other Asian and African countries.

## Data Availability

Data are available in the following GitHub repository - https://github.com/indrajitg-r/COMET-LF.

https://www.indiastatbihar.com/bihar-state/data/health/filaria

## Data and code availability

All the data used in the study are provided in the article. EPIFIL and LYMFASIM codes are available at www.ntdmodelling.org/diseases/lymphatic-filariasis.

## Acknowledgement

The authors would like to thank Dr. Bhupendra Tripathi and Dr. Luc Coffeng for helpful suggestions. We thank Dr. Nabanita Majumder for providing estimates of the migration rate. The work/opinion is based on research findings by the authors and do not reflect the opinion of the government.

## Author Contributions

**Conceptualization:** Subramanian Swaminathan, Souvik Banerjee, Mithun K. Mitra.

**Data Curation:** Indrajit Ghosh, Suchita Nath-Sain.

**Formal Analysis:** Indrajit Ghosh.

**Funding Acquisition:** Souvik Banerjee, Mithun K. Mitra.

**Investigation:** Indrajit Ghosh, Suchita Nath-Sain.

**Methodology:** Indrajit Ghosh, Subramanian Swaminathan, Souvik Banerjee, Mithun K. Mitra.

**Software:** Indrajit Ghosh, Shoummo Sengupta, Subramanian Swaminathan

**Supervision:** Chhavi Pant Joshi, Tanu Jain, Souvik Banerjee, Mithun K. Mitra.

**Visualization:** Indrajit Ghosh, Suchita Nath-Sain, Mithun K. Mitra.

**Writing – Original Draft:** Indrajit Ghosh, Suchita Nath-Sain.

**Writing – Review & Editing:** Indrajit Ghosh, Suchita Nath-Sain, Shoummo Sengupta, Subramanian Swaminathan, Chhavi Pant Joshi, Tanu Jain, Souvik Banerjee, Mithun K. Mitra.

